# Whole Exome Sequencing Analyses Support a Role of Vitamin D Metabolism in Ischemic Stroke

**DOI:** 10.1101/2022.05.31.22275825

**Authors:** Yuhan Xie, Julián N. Acosta, Yixuan Ye, Zachariah S. Demarais, Carolyn J. Conlon, Ming Chen, Hongyu Zhao, Guido J. Falcone

## Abstract

Ischemic stroke (IS) is a highly heritable trait. Genome-wide association studies have identified several commonly occurring susceptibility risk loci for this condition. However, there are limited data on the contribution of rare genetic variation to IS. We conducted a whole-exome association study of IS in 152,058 UK Biobank participants (mean age 57, 6.8 [SD 8.0], 83,131 [54.7%] were females), including 1,777 IS cases (mean age 61.4 [SD 6.6], 666 [37.5%] were females). We performed single-variant analyses for all variants and gene-based analyses for loss of function and deleterious missense rare variants. In the gene-based analysis, rare genetic variation at *CYP2R1* was significantly associated with IS risk (*P*=2.6×10^−6^), exceeding the Bonferroni-corrected threshold for 16,074 tests (*P*<3.1 × 10^−6^). We first replicated these findings using summary statistics from a genome-wide association study that included 67,162 IS cases and 454,450 controls (gene-based test for *CYP2R1, P*=0.003). We pursued a second replication focused on IS recurrence using individual-level data from 1,706 IS survivors, including 142 cases of recurrent IS, enrolled in the VISP trial (gene-based test for *CYP2R1, P*=0.001). We also found that common genetic variation at *CYP2R1* was associated with white matter hyperintensity volume (42,310 participants) and both mean diffusivity and fractional anisotropy (17,663 participants) in the subcohort of UK Biobank (all gene-based tests *P*<0.05). Because *CYP2R1* plays an important role in vitamin D metabolism, our results support a role of this pathway in the occurrence of ischemic cerebrovascular disease.

## Introduction

Ischemic stroke is one of the leading causes of death and disability worldwide.^1^ In addition, mounting evidence indicates that ischemic stroke significantly contributes to cognitive decline and dementia.^2,3^ Despite significant advances in stroke prevention and treatment, a substantial proportion of ischemic stroke patients are not eligible for reperfusion therapies.^4^ Importantly, the overall impact of ischemic stroke seems to be significantly modified by minority status^5–8^ and is much higher in low and middle income countries.^1,9^

Mounting evidence indicates that common genetic variation substantially contributes to the occurrence of ischemic stroke. This genetic contribution influences not only risk, but also the severity, outcome and recurrence of stroke.^10^ The largest published genome wide association study (GWAS) of ischemic stroke to date identified 32 susceptibility risk loci that modify all stroke subtypes, including cardioembolic, small vessel, and large artery stroke.^11^ In addition, follow-up studies have estimated the heritability of ischemic stroke at ∼38%.^12^ However, due to the lack of large studies with available sequencing data, most efforts to date have focused on common and low-frequency genetic variants, with only a few addressing the contribution of rare variants.

Leveraging the availability of whole exome sequencing (WES) data from 200,632 participants enrolled in the UK Biobank,^13^ we aimed to identify rare genetic risk factors for ischemic stroke.

## Materials and methods

### Study design

We conducted a genetic association study using whole exome sequencing and clinical data from the UK Biobank. The UK Biobank is a population-based study that enrolled 502,618 participants across the United Kingdom. Ethics approval for the study was obtained from the North West Multi-centre Research Ethics Committee as a Research Tissue Bank approval. Participants in this study provided electronic signed consent, filled questionnaires on socio-demographic, lifestyle, health related factors, completed physical measures and medical tests, and provide DNA samples for future analysis. For this study, we included 200,632 study participants with available WES data, GWAS data and clinical data under project application number 29900.

### DNA sampling and genomic data

DNA was extracted from stored blood samples that had been collected at the time of enrollment and genotyping was carried out by the Affymetrix Research Services Laboratory. Genome-wide array genotyping was completed using the Applied Biosystems UK BiLEVE Axiom Array which contains 807,411 markers for a subset of 49,960 participants, and the closely related Applied Biosystems UK Biobank Axiom Array which contains 825,927 markers (95% of which were shared with UK BiLEVE Axiom Array) for 438,427 participants. Standard quality control procedures for genome-wide data were performed centrally by the UK Biobank research team including genotype-level filters, subject-level quality checks, and imputation. The pipeline yielded a final result of 93,095,623 autosomal SNPs, short indels, and large structural variants in 487,442 individuals. A detailed description of DNA sampling, genotyping, and quality control procedures were described elsewhere.^14^ The BGEN version of genome-wide genotyping data for the result was downloaded from the website of UK Biobank.

Exomes were captured using the IDT xGen Exome Research Panel v1.0 including supplemental probes^15^. The samples were sequenced with dual-indexed 75× 75 bp paired-end reads on the Illumina NovaSequence 6000 platform. The first ∼50,000 tranche was sequenced with IDT v1.0 oligo lot using S2 flow cells, and the subsequent ∼150,000 samples were processed with the 150,000 oilgo lot using S4 flow cells. The PLINK version of whole exome sequencing data under OQFE protocol for 200,632 individuals was downloaded from the website of UK Biobank. More information on the WES OQFE protocol was introduced in the original release paper.^13^

### Outcome ascertainment

The present study focuses on IS, the most frequent stroke subtype. To maximize power, we included both prevalent (present upon enrollment) and incident (identified during follow-up) IS cases. IS cases were identified using a previously validated algorithm that integrates International Classification of Diseases (ICD) codes, self-reported data, and information from national death registries. Definitions for baseline characteristics are provided in Supplementary Table 1.

### Vitamin D levels

The UK Biobank measured 36 biochemistry markers in all 500,000 study participants, including serum Vitamin D level. These were measured using the Chemiluminescence ImmunoAssay test through LIASON XL by DiaSorin Ltd. A series of robust and detailed quality procedures were employed to minimize and mitigate the effects of systematic bias and random error.^16^ The phenotype data of serum Vitamin D level were acquired from UK Biobank data field ID 30890.

### Quality control

We only kept variants from autosomal chromosomes in our analyses. Variants with missingness greater than 10% and Hardy-Weinberg equilibrium less than 1×10^−6^ were removed. We defined the relatedness between samples using the kinship coefficient score (KING). For samples with KING>0.0442, we first removed related samples who had more than or equal to two relatives and iteratively removed those individuals until none remained. Then for each pair of remaining related individuals, we only removed a single sample at random. A subset of European ancestry samples was selected based on the UK Biobank data field ID 22006. Participants who did not have IS but had other stroke subtypes were further removed from the samples.

### Variant-level functional annotations

We annotated the retained variants in our study cohort with minor allele frequencies (MAF) and functional categories using the genome build GRCh38. Loss of function (LoF) variants were those predicted to cause frameshift insertion/deletion, splice site alteration, stop gain, and stop loss, and deleterious missense (Dmis) variants were defined as those predicted consistently to be deleterious by 9 in silico prediction including SIFT^17^, Polyphen2_HVAR^18^, Polyphen2_HDIV^18^, M-CAP^18,19^, MetaLR^20^, MetaSVM^20^, LRT^21^, PROVEAN^22^, and MutationTaster^22,23^.

### Single variant association analysis

We conducted single variant analysis for rare variants with MAF<0.01 using the software package REGENIE. For step 1, we used the genotype array data from UK Biobank to fit the linear mixed model and followed the quality control steps in their documentation. For step 2, we applied the approximate firth logistic regression, used 400 as the block size and the default settings for other parameters. Sex, age, age^2^, the interaction of age and sex, and the first 4 genomic principal components were adjusted as covariates in the analyses. Genome-wide significance was set as 5×10^−8^ in our analysis.

### Gene-based rare variant association analysis

We only included rare LoF and Dmis variants with MAF<0.01 in our gene-based association test. Furthermore, LoF and Dmis variants were collapsed as the damaging variants and included in our gene-based test. We applied the robust method of the Optimal Unified Test (SKAT-O)^24^ to conduct the gene-based test. Further, ultra-rare variants with MAF>1×10^−5^ were removed to mitigate the potential inflation due to low minor allele counts.^25^ The same set of covariates as the single variant analysis was adjusted in the gene-based test. Exome-level significance was set as 0.05 over the total number of genes tested.

### Replication using individual-level data

We replicated observed associations using individual-level data from the Genomics and Randomized Trials Network (GARNET), a genetic substudy of the Vitamin Intervention for Stroke Prevention (VISP) clinical trial. Detailed descriptions of GARNET and VISP can be found elsewhere.^26^ Briefly, VISP evaluated whether high doses of folic acid, pyridoxine (vitamin B6), and cobalamin (vitamin B12), given to lower total homocysteine levels, reduce the risk of recurrent stroke over a 2-year period compared with low doses of these vitamins. GARNET genotyped a portion of the study participants enrolled in the parent clinical trial using the Illumina HumanOmni1-Quad-v1 array (Illumina, Inc.). Phenotypic and genotypic data from GARNET were acquired through the database of Genotypes and Phenotypes (dbGaP) (accession number phs000343.v3.p1). Genome-wide data were quality controlled using standard filters, population structure was accounted for via multidimensional scaling,^27^ and data were pre-phased and imputed to 1000 Genomes integrated reference panels (phase 3 integrated variant set release in NCBI build 37).^28^ A detailed preprocessing procedure are presented in Supplementary Table 2.

We used logistic regression to perform single-variant association tests with recurrent stroke, adjusting for the treatment group (i.e., high-dose vs low-dose vitamin supplementation), sex, age, age^2^, the interaction of age and sex, and the first 4 genomic principal components. Variant-level significance was set as 0.05 over the total number of SNPs tested. We used the GATES^29^ test to also conduct gene-based tests.

### Replication using summary statistics

We performed replication analyses using summary results from MEGASTROKE^11,30^ (the largest GWAS of all stroke types conducted to date), a recent GWAS of small vessel stroke^31^, a recent GWAS of MRI markers for cerebral small vessel disease in UK Biobank,^32^ and the Biobank Japan.^33,34^ We downloaded the summary statistics from the Cerebrovascular Disease Knowledge Portal.^35^ Summary statistics of markers within the 50kb of our targeting gene and linkage disequilibrium (LD) structure from reference panels were set as input for the GATES test^29^. To model the LD structure, we used European and East Asian panels from the 1000 Genomes Project. SNPs with INFO score<0.8, MAF<0.001, and deviated from Hardy-Weinberg equilibrium (*P*<1×10^−10^) were removed. We only considered the overlapping markers between the GWAS summary statistics and the LD reference panel in the GATES tests.

### Software and packages

For UK Biobank WES, quality control on variants was conducted with PLINK 1.90^27^ and quality control on individuals was conducted with PLINK 1.90 and R 3.5.0 Variant-level MAF was annotated by PLINK 1.90, and functional categories of variants were annotated by ANNOVAR^36^. For the gene-based association test, we applied the robust function implemented in the R package SKAT that can deal with unbalanced cases and controls^24^. Manhattan plots were generated using an adapted function from the R package qqman^37^. For GARNET, quality control was conducted with PLINK 1.90, SHAPEIT^38^ and IMPUTE2^39^. Regional GWAS results were visualized using LocusZoom^40^. For gene-based analysis, we applied the GATES^29^ test implemented in the R package aSPU.

## Results

Out of a total of 502,618 study participants enrolled in the UK Biobank, 200,632 underwent whole exome sequencing. Of these, 16,335 were removed from the analysis when applying subject-level filters, including 16,121 for relatedness and 214 for sex mismatch or aneuploidy. Within this population of study participants who passed subject-level quality control filters, 152,710 were of European ancestry. Among the European ancestry subset, 1,777 individuals developed an ischemic stroke. 652 individuals who had sustained hemorrhagic strokes were removed from the analysis. A total of 152,058 participants (mean age 56.8, female sex 83,141) were included in our analysis. Out of a total of 17,549,650 genotyped genetic variants, 428,399 were removed from the analysis when applying variant-level filters, including 289,101 for missingness and 139,298 for Hardy-Weinberg equilibrium (Table 1).

**Table 1.**
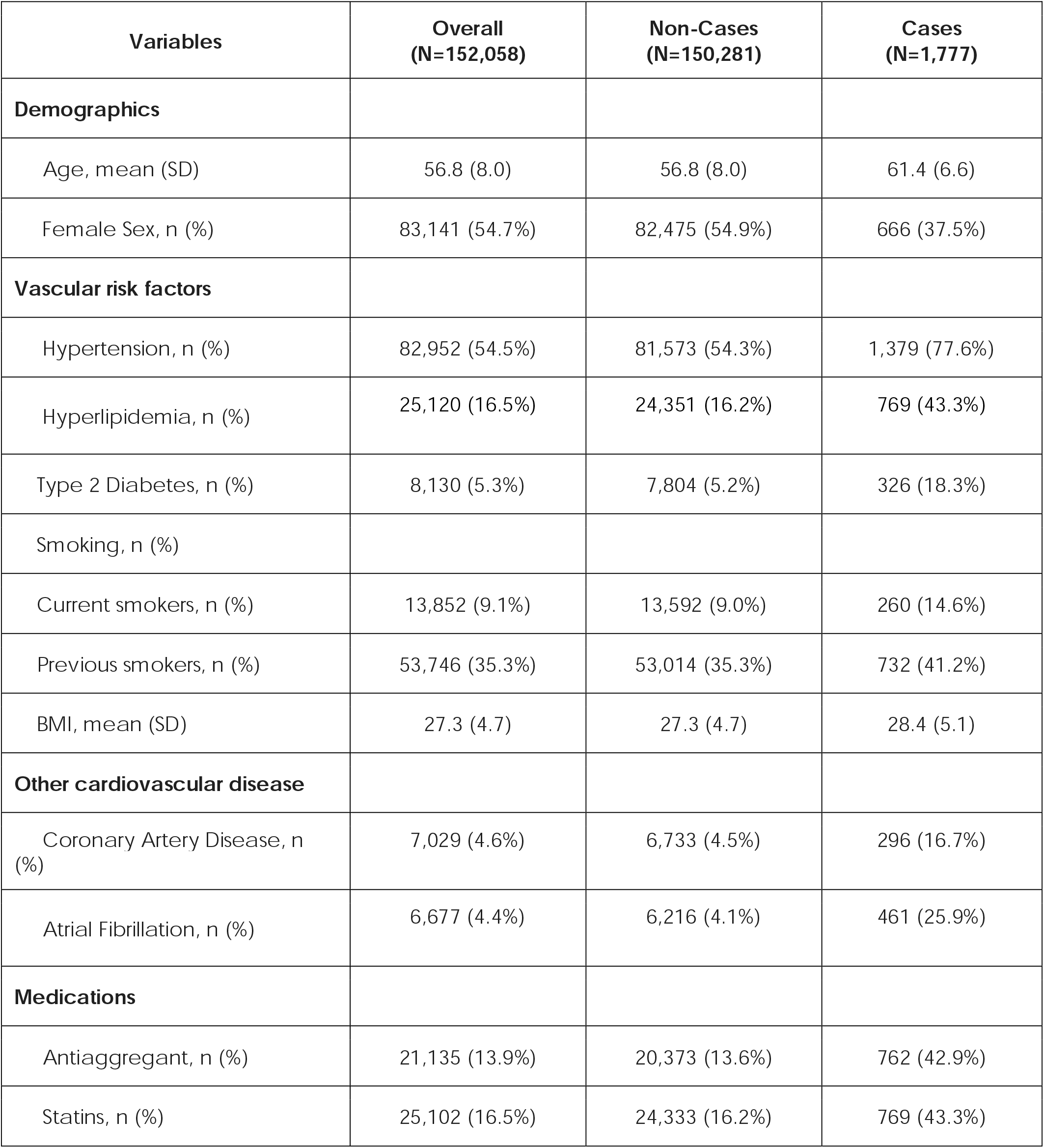
Baseline characteristics of the study population.

| **Variables** | **Overall**<br>**(N=152,058)** | **Non-Cases**<br>**(N=150,281)** | **Cases**<br>**(N=1,777)** |
| --- | --- | --- | --- |
| **Demographics** |  |  |  |
| Age, mean (SD) | 56.8 (8.0) | 56.8 (8.0) | 61.4 (6.6) |
| Female Sex, n (%) | 83,141 (54.7%) | 82,475 (54.9%) | 666 (37.5%) |
| **Vascular risk factors** |  |  |  |
| Hypertension, n (%) | 82,952 (54.5%) | 81,573 (54.3%) | 1,379 (77.6%) |
| Hyperlipidemia, n (%) | 25,120 (16.5%) | 24,351 (16.2%) | 769 (43.3%) |
| Type 2 Diabetes, n (%) | 8,130 (5.3%) | 7,804 (5.2%) | 326 (18.3%) |
| Smoking, n (%) |  |  |  |
| Current smokers, n (%) | 13,852 (9.1%) | 13,592 (9.0%) | 260 (14.6%) |
| Previous smokers, n (%) | 53,746 (35.3%) | 53,014 (35.3%) | 732 (41.2%) |
| BMI, mean (SD) | 27.3 (4.7) | 27.3 (4.7) | 28.4 (5.1) |
| **Other cardiovascular disease** |  |  |  |
| Coronary Artery Disease, n (%) | 7,029 (4.6%) | 6,733 (4.5%) | 296 (16.7%) |
| Atrial Fibrillation, n (%) | 6,677 (4.4%) | 6,216 (4.1%) | 461 (25.9%) |
| **Medications** |  |  |  |
| Antiaggregant, n (%) | 21,135 (13.9%) | 20,373 (13.6%) | 762 (42.9%) |
| Statins, n (%) | 25,102 (16.5%) | 24,333 (16.2%) | 769 (43.3%) |

### Single Variant Association Analysis

In the exome-wide, single variant association analysis none of the evaluated genetic variants were significantly associated with the risk of ischemic stroke at the genome-wide significance level (*P*□<□5 × 10^−8^). There was one noncoding variant on gene *ANK2* at the margin of significance (*P*=7.9×10^−8^; 0R=0.02, *SE*=1.84).

### Gene-Based Rare Variant Association Analysis

We performed a gene-based, exome-wide association analysis using the SKAT-O robust method implemented in the SKAT software. The Manhattan plot and Quantile-Quantile (Q-Q) plot for a total number of 16,074 genes tested across the human genome are presented in Figure 1. *CYP2R1* was the only gene significantly associated with ischemic stroke (*P*=2.6 × 10^−6^, surpassing the Bonferroni-corrected threshold of 3.1×10^−6^, adjusted for 16,074 tests). In total, there were 21 LoF and Dmis variants included in the test for *CYP2R1*. Two additional genes *CCDC74B* (*P*=2.3×10^−5^) and *PLOD2* (*P*=6.1×10^−5^) showed suggestive associations (Figure 1).

**Figure 1.**
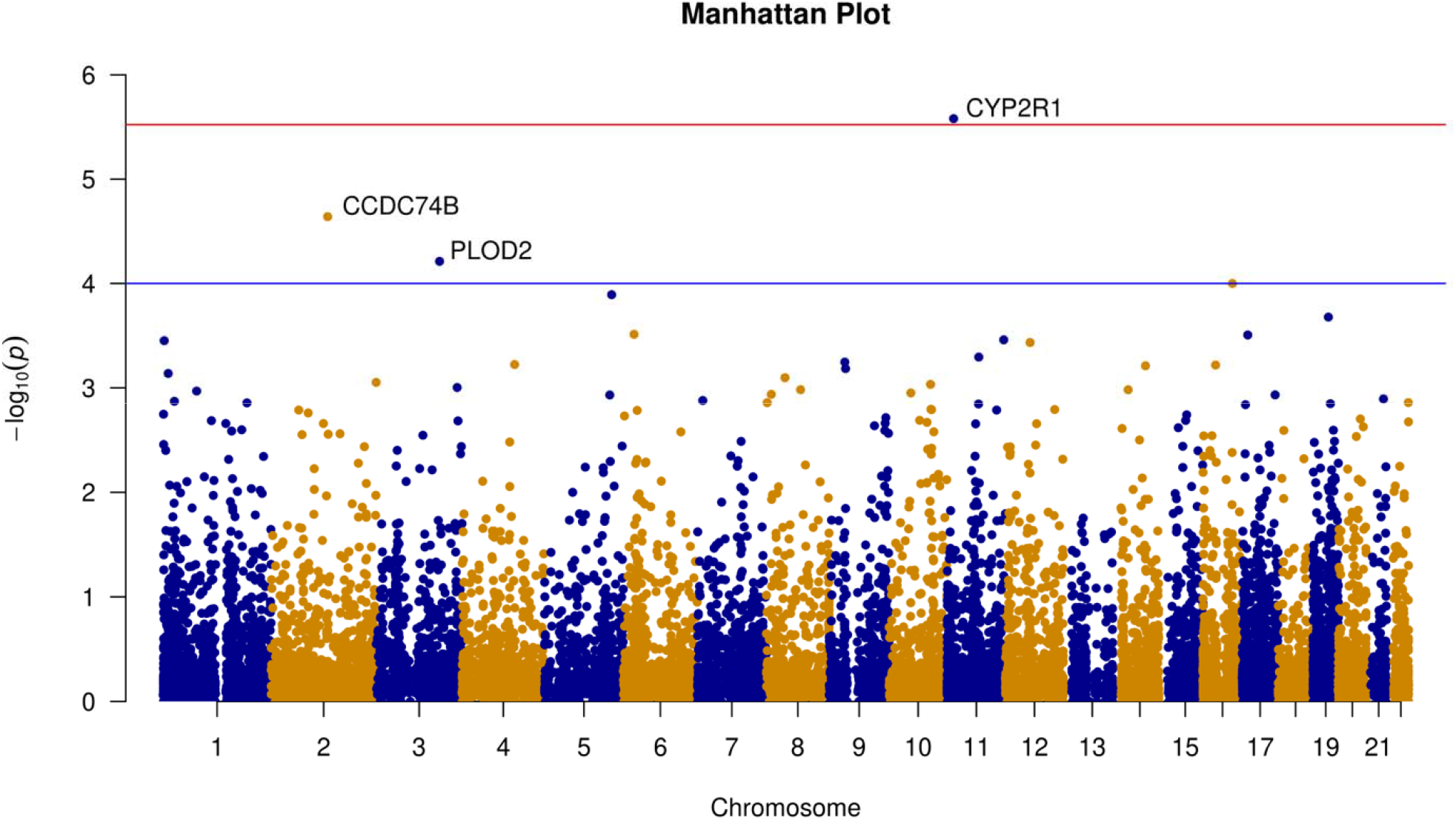
Manhattan plot for the exome-wide gene-based association analysis. The genome coordinates of each variant are displayed along the x-axis, and the negative logarithm of the association p-values for each variant are displayed on the y-axis. The blue line represents the threshold of, and the red line represents the threshold of. Genes that passed the thresholds in our gene-based tests are annotated with gene symbols.

Given the central role of *CYP2R1* in the metabolism of vitamin D, we tested for association between rare genetic variation at *CYP2R1* and serum levels of vitamin D in study participants with available measurements (139,015 participants), finding a highly significant association (*P*=1.3×10^−10^). Next, in this same group of patients with available Vitamin D measurements, we investigated serum levels of vitamin D in all participants, participants with any rare *CYP2R2* variants, participants with rare *CYP2R2* variants not observed in stroke patients, and participants with rare *CYP2R2* variants observed in stroke patients. The median concentration of serum levels of vitamin D in these 4 groups was 48.2, 38.0, 39.7 and 35.9 (*P*<0.001Table 2 and Figure 2).

**Table 2.** Mean and median vitamin D levels in all studied participants and carriers of rare *CYP2R1* variants.

| Evaluated group | Number of Participants | Mean | Median |
| --- | --- | --- | --- |
| All | 139,015 | 49.7 | 48.2 |
| Carriers of any rare variants | 181 | 40.1 | 38.0 |
| Carriers of variants observed in stroke patients | 54 | 37.8 | 35.9 |
| Carriers of other rare variants | 127 | 41.0 | 39.7 |

**Figure 2.**
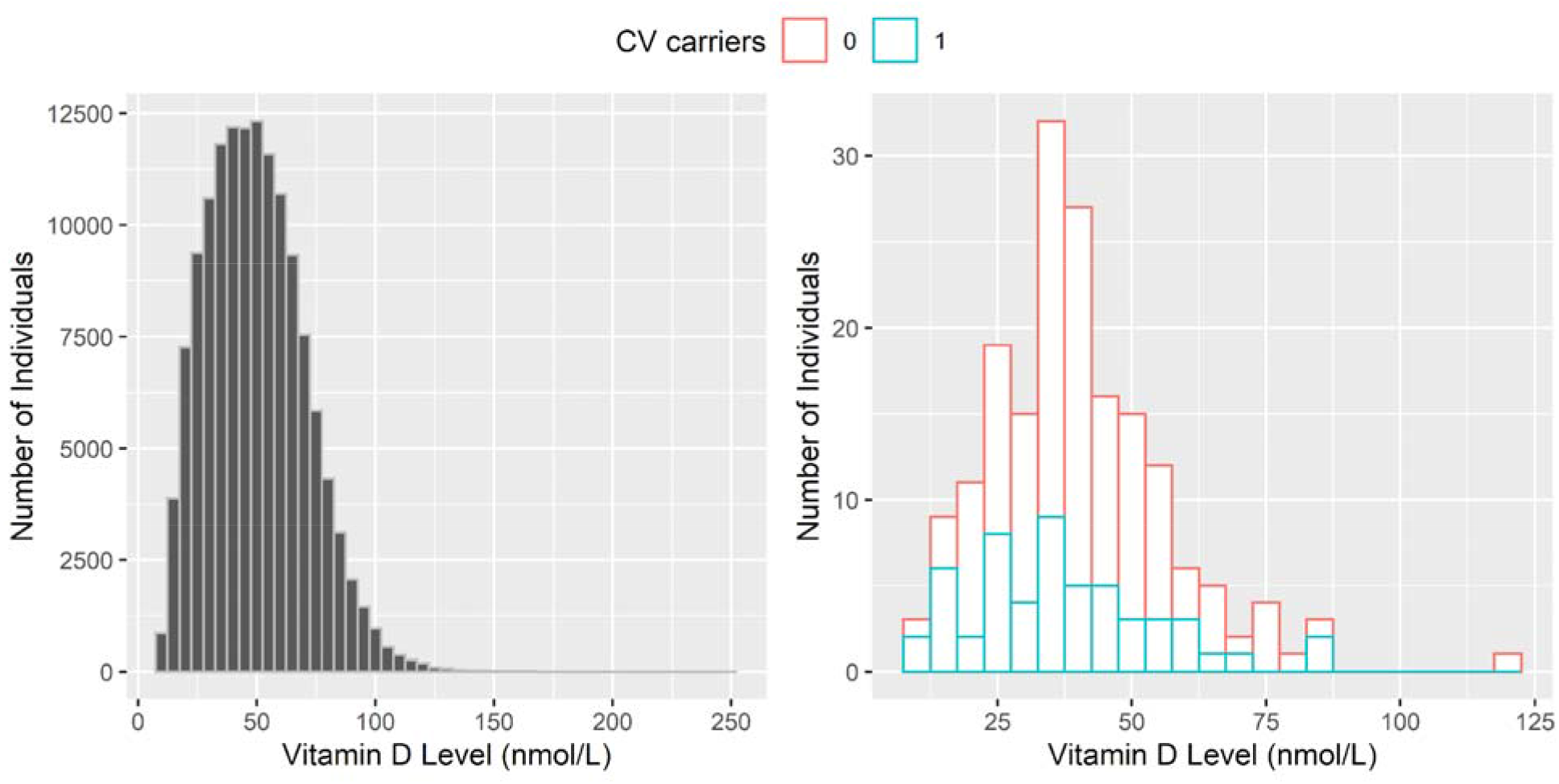
Vitamin D levels in all participants and *CYP2R1* carriers. **(A)** Histogram of serum vitamin D levels in all participants included in the analysis. **(B)** Histogram of serum Vitamin D levels in carriers of *CYP2R1* variants; in red non-CV carriers, in blue CV carriers.

**Figure 3.**
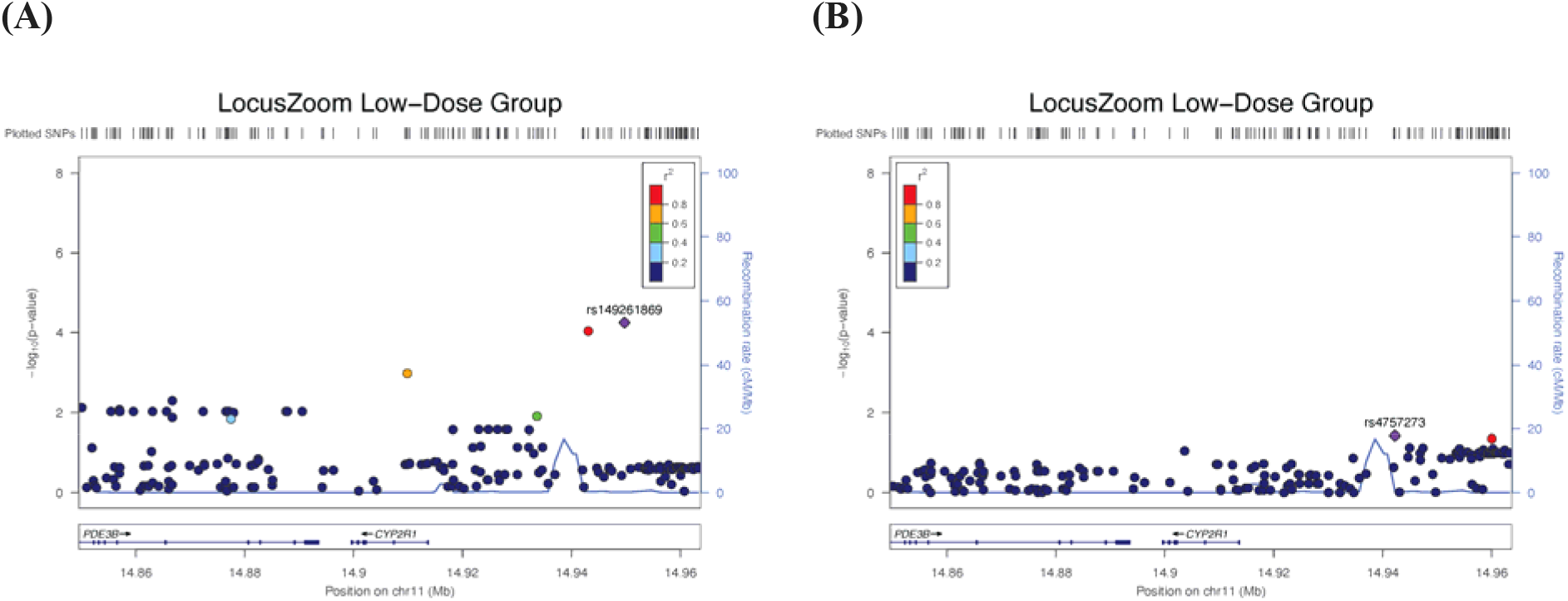
Locus Zoom plots of the association analysis of *CYP2R1* variants in GARNET by treatment arm. The genome coordinates of each variant are displayed along the x-axis, and the negative logarithm of the association p-values for each variant are displayed on the y-axis. **(A)** Low-dose group. **(B)** High-dose group.

### Replication in the GARNET study using individual-level data

We tested for association between *CYP2R1* and recurrent stroke using both gene-based tests and single variant analysis. When implementing gene-based testing, we replicated the association between *CYP2R1* and ischemic stroke in the entire cohort (171 evaluated variants, *P*=1.2×10^−3^), the subgroup randomized to low-dose vitamin supplementation (161 evaluated variants, *P*=6.0×10^−5^), and the subgroup randomized to high-dose vitamin supplementation (166 evaluated markers, *P*=0.04). When implementing single variant analyses, the makers rs149261869 (*P*=5.6 × 10^−5^) and rs117335120 (*P*=9.2 × 10^−5^) located near a recombination region were significantly associated with the risk of ischemic stroke in the subgroup randomized to low-dose vitamin supplementation.

### Replication using summary statistics

We conducted gene-based testing using summary statistics from MEGASTROKE, the largest GWAS of ischemic stroke completed to date, separately evaluating all stroke types combined (labeled any stroke), any ischemic stroke, large artery stroke, cardioembolic stroke, and small vessel stroke (SVS). We also completed similar gene-based tests using summary statistics from a recent, large GWAS focused on SVS, GWAS of neuroimaging traits related to clinically silent cerebrovascular disease (including white matter hyperintensities volume, fractional anisotropy, and mean diffusivity), and GWAS summary statistics for ischemic stroke in Biobank Japan. All gene-based test p-values except for MEGASTROKE SVS (*P*=0.07) were significant at the significance level of 0.05 (Table 3).

**Table 3.** Results of replication analyses using summary statistics from genome-wide association studies and meta-analysis from several ischemic stroke related phenotypes.

| Study / Phenotype | Total Markers | # of Markers Tested | P-Value |
| --- | --- | --- | --- |
| MEGASTROKE<br>Any stroke (ischemic or hemorrhagic) | 8,255,860 | 158 | 0.01 |
| MEGASTROKE<br>Any ischemic stroke | 8,340,184 | 158 | $3.6 \times 10^{-3}$ |
| MEGASTROKE<br>Large artery ischemic stroke | 8,451,005 | 162 | 0.003 |
| MEGASTROKE<br>Cardioembolic ischemic stroke | 8,306,090 | 158 | $3.6 \times 10^{-3}$ |
| MEGASTROKE<br>Small vessel ischemic stroke | 8,765,828 | 159 | 0.08 |
| GWAS of small vessel ischemic stroke | 6,939,580 | 144 | $3.2 \times 10^{-3}$ |
| GWAS of white matter hyperintensities | 9,733,965 | 196 | 0.03 |
| GWAS of DTI metrics<br>Fractional anisotropy | 9,735,097 | 196 | 0.004 |
| GWAS of DTI metrics<br>Mean diffusivity | 9,735,314 | 195 | 0.05 |
| GWAS of ischemic stroke<br>Biobank Japan | 8,678,632 | 176 | 0.02 |

## Discussion

In this study, we leveraged data from the UK Biobank to conduct an exome-wide association analysis of ischemic stroke using both single-variant and gene-based testing. The gene-based analyses revealed that rare genetic variation at *CYP2R1* was associated with a higher risk of ischemic stroke. This gene codes for the cytochrome P450 2R1, the enzyme in charge of the first step of the activation of vitamin D. We therefore showed that rare genetic variation at *CYP2R1* was strongly associated with lower levels of circulating vitamin D. Importantly, these results were replicated using both individual-level data and summary statistics from several different studies of stroke and other phenotypes related to cerebrovascular diseases.

It has been widely demonstrated that common genetic variation contributes to stroke risk and severity.^10^ The largest GWAS of ischemic stroke conducted to date identified 32 susceptibility risk loci for the different types of ischemic stroke that are seen in clinical practice. In addition, other published GWAS focused on functional outcome and pre-clinical studies using animal models also indicate that common and rare genetic variation contributes to stroke recovery as well.^41,42^ Beyond ischemic stroke, common genetic variation also influences intraparenchymal hemorrhage and subarachnoid hemorrhage, the most common subtypes of hemorrhagic stroke.^43,44^

Despite the compelling evidence for the role of genetic variation in stroke risk and outcome, most studies to date have focused on common genetic variation, with only a few studies investigating the role of rare variants. A recent large study from the Trans-Omics Precision Medicine (TOPMed) program conducted a whole genome association analysis and found four susceptibility loci driven by rare variants. One of these loci was specifically related to large artery ischemic stroke (13q33, top associated variant rs181401679), two were associated with stroke of any mechanism (*RAP1GAP2*-rs60380775 and *AUTS2*-rs150022429) and the fourth was associated with hemorrhagic stroke (7q22-rs141857337).^45^ Additionally, the study found one locus (*TEX13C*-rs145400922) associated with cardioembolic stroke at the genome-wide significance level in analysis restricted to Black participants. However, replication is still needed for these results, and sample size represents a challenge given most of these variants are rare. Further, a whole-exome-wide analysis of white matter hyperintensities burden, a neuroimaging trait that represented clinically silent cerebrovascular disease, identified one susceptibility risk locus driven by rare genetic variation at *HTRA1*. Of note, carriers of rare mutations in this gene showed a larger effect than clinical risk factors such as hypertension, diabetes, obesity, and smoking.^46^

Our study provides significant new evidence to the field of stroke genomics research focused on rare genetic variation. In an exome-wide analysis using gene-based testing, we identified *CYP2R1* as a novel susceptibility risk locus for ischemic stroke. *CYP2R1* is located at 11p15.2 and encodes the cytochrome P450 2R1, a vitamin D 25-hydroxylase involved in the first step of the activation of vitamin D.^47,48^ Of note, a previous study demonstrated that one low-frequency variant in this gene leads to a 2-fold increase in risk of vitamin D insufficiency,^49^ in line with our results showing that carriers of *CYP2R1* variants had, on average, lower vitamin D levels in the UK Biobank. Importantly, we pursued replication of these findings using summary statistics from several well-powered GWAS of ischemic stroke (including several clinically relevant subtypes) and neuroimaging traits of clinically silent cerebrovascular disease,^50^ with all but one of these analyses achieving nominal significance. Furthermore, we also replicated the association between *CYP2R1* and ischemic stroke using gene-based testing and individual-level data from GARNET, a genetic sub study of the VISP clinical trial, that evaluated the role of high versus low multi-vitamin supplementation in stroke recurrence after a first stroke.

Many observational, experimental, and genetic studies have investigated the role of vitamin D on cardiovascular endpoints.^51,52^ While many observational studies have suggested a relationship between lower vitamin D levels and worse cardiovascular health,^53–57^ most randomized clinical trials and Mendelian randomization studies have failed to confirm a causal effect.^58–64^ However, many of these studies assumed a linear relationship between vitamin D levels and the diseases of interest, and recent studies suggest that a J-shaped relationship is more likely to accurately reflect the biological relationship between the metabolism of vitamin D and cardiovascular disease. Along these lines, a recent observational and genetic analysis of vitamin D levels and a wide range of clinical endpoints demonstrated a significant causal association between vitamin D and all-cause mortality in patients with vitamin D deficiency.^65^ Similarly, a recent review concluded that vitamin D supplementation likely carries no benefit in persons with normal levels of this vitamin but advocated for routine supplementation in persons with low levels.^66^

In future steps, additional replication of our findings in other cohorts is warranted. Specifically, analysis of WES data from individuals from other ancestries is urgently needed, as our study only included participants of European ancestry. Further steps include the analysis of gene-environment interactions that could play a role in ischemic stroke risk. In addition, we only tested the association between genes and stroke risk, but several studies have highlighted that many exposures that influence risk also modify disease severity and trajectories in patients with stroke. In that context, additional studies exploring the role of *CYP2R1* in stroke severity, outcome, and recurrence are needed. Of note, given the huge sample sizes needed to obtain enough statistical power to identify rare genetic risk factors,^67^ the importance of large international consortia and collaborations is paramount.

Our study has several limitations. First, replication of our results using individual-level data from WES/WGS studies is still needed. Second, ischemic stroke is a heterogeneous condition with many subtypes that share an underlying biology and risk factors, but also have distinct features.^68^ Unfortunately, information on ischemic stroke subtypes in the UK Biobank is not available, and therefore these subtypes could not be analyzed separately. Finally, the ascertainment of ischemic stroke in the UK Biobank is based on self-reported, EHR-based, and death reports data, which could introduce inaccuracies. However, most ascertainment was done based on EHR validated codes, which has shown to have a positive predictive value of 80-90%.^69,70^

In summary, we leveraged WES data from 152,058 participants from the UK Biobank and found that *CYP2R1*, a gene encoding the vitamin D 25-hydroxylase, a key enzyme for vitamin D activation, is associated with ischemic stroke risk in persons from European ancestry. Future research to validate our findings in other cohorts, especially including participants from other ancestries, is warranted.

## Data Availability

Genotype and phenotype data from UK Biobank (www.ukbiobank.ac.uk) and GARNET (https://www.ncbi.nlm.nih.gov/projects/gap/cgi-bin/study.cgi?study_id=phs000343.v4.p1) cannot be shared publicly because only approved users can have access. GWAS summary data used in this study are publicly accessible (https://www.kp4cd.org/dataset_downloads/stroke).

## Acknowledgments

We conducted the research using the UK Biobank resource under an approved data request (ref: 29900). We sincerely thank the UK Biobank and its participants for their contribution to science and many genome-wide association studies (GWAS) consortia for making their GWAS summary data publicly accessible (https://www.kp4cd.org/dataset_downloads/stroke).

## Author contributions

Conception and design of the study: YX, JNA, HZ, GJF

Acquisition and analysis of the data: YX, JNA, HZ, GJF, YY, MC, ZSD, CJC

Drafting of the manuscript and preparation of tables: YX, JNA, HZ, GJF, ZSD, CJC

## Supplementary material

Supplementary Table 1

Supplementary Table 2

## Abbreviations

IS: ischemic stroke
GWAS: genome wide association study
WES: whole exome sequencing
KING: kinship coefficient score
LD: linkage disequilibrium
MAC: minor allele count
MAF: minor allele frequencies
LoF: loss of function
Dmis: deleterious missense
SVS: small vessel stroke
TOPMed: Trans-Omics Precision Medicine
dbGaP: database of Genotypes and Phenotypes
GARNET: Genomics and Randomized Trials Network
VISP: Vitamin Intervention for Stroke Prevention

