## Supplemental material for "Whole Exome Sequencing Analyses Support a Role of Vitamin D Metabolism in Ischemic Stroke"

Supplementary Data

### Supplementary Table 1. Baseline Characteristics in the UK Biobank.

|  |  | **Field IDs** | **Criteria** | **ICD-9** | **ICD-10** | **OPCS-4** |
| --- | --- | --- | --- | --- | --- | --- |
| **Demographics** | Age | 21022 | - | - | - | - |
|  | Sex | 31 | - | - | - | - |
| **Vascular Risk Factors** | Hypertension | 4079,4080 | Systolic blood pressure>140 mmHg or  Diastolic blood pressure>90 mmHg | - | - | - |
|  | Hyperlipidemia | 20003  23400 | Use Statins (see criteria below) or  Total Cholesterol>200mg/dl | - | - | - |
|  | Type 2 Diabetes | 20002 (1223) | - | - | E11 | - |
|  | Smoking | 20116 | - | - | - | - |
|  | BMI | 21001 | - | - | - | - |
| **Co-morbidities** | Coronary Artery Disease | 20002 (1075)  20004(1070,1095) | - | 410-412 | I21-I23, I241, I252 | K40.1-40.4, K41.1-41.4, K45.1-45.5, K49.1-49.2, K49.8-49.9, K50.2, K75.1-75.4, K75.8-75.9 |
|  | Atrial Fibrillation | 20002(1471,1483)  20004 (1524) | - | 427.3 | I48 | K57.1, K62.1-K62.4 |
| **Medications** | Antiaggregant | 20003 | Aspirin, clopidogrel | - | - | - |
|  | Statins | 20003 | Rosuvastatin, simvastatin, fluvastatin, pravastatin, atorvastatin | - | - | - |

### Supplementary Table 1: GARNET Dataset Quality Control

| **GENOTYPING** |  |
| --- | --- |
| Genotyping Center | Center for Inherited Disease Research, Johns Hopkins University |
| Genotyping Array | Illumina HumanOmni1-Quad-v1 array (Illumina, Inc.) |
| **SAMPLE QC** |  |
| **Starting cohort** | **2,122** |
| Samples with call rate > 0.95 | 2,122 (-0) |
| No sex mismatch | 2,108 (-14) |
| Not heterozygosity outlier | 2,103 (-5) |
| Relatedness IBD < 0.2 | 2,103 (-0) |
| Population stratification European (via MDS) | 1,761 (-342) |
| Retrieving chromosome 11 | 1,758 (-3) |
| Retrieving samples labelled “white” | 1,706 (-52) |
| **For analysis** | **1,706** |
| **SNP QC** |  |
| Biallelic SNPs | 1,140,419 |
| SNPs with call rate > 0.95 | 997,504 (-142,915) |
| Autosomal SNPs | 972,412 (-25,092) |
| SNPs with minor allele frequency >0.01 | 867,043 (-105,369) |
| SNPs not deviating from HWE (<10^−6^) | 800,742 (-66,301) |
| SNPs with non-sig different call rate cases vs controls (p < 0.05) | 791,723 (-9,019) |

QC = Quality control, SNP = Single nucleotide polymorphism, HWE = Hardy-Weinberg equilibrium, IBD = Identity-by-descent, MDS = Multidimensional Scaling
